# Risk Assessment and Predictive Modeling of Suicide in Chronic Lymphocytic Leukemia/Small Lymphocytic Lymphoma (CLL/SLL) Patients

**DOI:** 10.1101/2025.04.15.25325875

**Authors:** Fan Wang

**Affiliations:** Department of Hematology, Tongji Hospital, Tongji Medical College, Huazhong University of Science and Technology, Wuhan, Hubei, The People’s Republic of China

**Keywords:** CLL/SLL, SEER, suicide risk, predictive modeling, nomogram

## Abstract

**Background:** Chronic lymphocytic leukemia/small lymphocytic lymphoma (CLL/SLL) predominantly affects older adults and is characterized by a prolonged disease course. While overall survival has improved, the psychosocial impact, including suicide risk, remains underexplored.

**Methods:** A retrospective cohort study was conducted using data from 95,517 patients diagnosed with CLL/SLL between 2000 and 2021, extracted from the SEER 17 registry. LASSO regression was utilized for variable selection, followed by univariate and multivariate Cox proportional hazards and Fine-Gray competing risk analyses to identify independent predictors. A nomogram was developed based on significant predictors and validated using time-dependent receiver operating characteristic (ROC) curves, calibration plots, and decision curve analysis (DCA).

**Results:** The cohort’s mean age was 69.2 years, with 21.6% aged ≥80 years and a male-to-female ratio of 1.5:1; 88.8% were White. Although the suicide rate was only 0.1%, multivariate analyses demonstrated that advanced age (≥80 years), male sex, single or non-married status, and lower median household income were significantly associated with increased suicide risk, while non-White race was associated with a lower risk. However, age lost statistical significance in the competing risk model. The nomogram demonstrated good discriminative ability, with area under the curve (AUC) values exceeding 0.71 at 3, 5, and 10 years in both training and validation cohorts. Calibration plots indicated good agreement between predicted and observed outcomes, and DCA confirmed clinical utility.

**Conclusions:** Sociodemographic factors, including sex, race, marital status, and income, are independently associated with suicide risk in CLL/SLL patients. The developed nomogram offers a practical, evidence-based tool for early identification of high-risk individuals, thereby facilitating targeted psychosocial interventions and improving survivorship care.

## Introduction

Chronic lymphocytic leukemia (CLL) and small lymphocytic lymphoma (SLL) are biologically identical, indolent B-cell neoplasms that differ primarily in anatomical distribution. CLL primarily involves the peripheral blood and bone marrow, while SLL is confined to lymphoid tissues^1^. As the most prevalent form of adult leukemia in Western populations, CLL predominantly affects older individuals, with a median age at diagnosis near 70 years^2^. Despite significant therapeutic advancements, particularly with the advent of targeted agents such as Bruton’s tyrosine kinase (BTK) inhibitors, CLL/SLL remains incurable and is characterized by a prolonged disease course, recurrent relapses, and the necessity for continuous clinical monitoring^3^. This extended disease trajectory often imposes a substantial psychological and emotional burden on affected individuals^4^.

An expanding body of evidence indicates that cancer patients are at significantly elevated risk of suicide compared to the general population^4^. Contributing factors include cancer-related psychological stress, depression, social isolation, functional decline, and financial strain^4,5^. Within hematologic malignancies, individuals diagnosed with leukemia or lymphoma consistently exhibit disproportionately high suicide rates^6^. For example, a large Scandinavian population-based study demonstrated that patients with hematologic cancers had nearly double the risk of suicide or attempted suicide within three years of diagnosis^5^. Similarly, an analysis of over 140,000 leukemia patients in the U.S. Surveillance, Epidemiology, and End Results (SEER) registry from 1975 to 2017 reported a suicide rate of 26.4 per 100,000 person-years, corresponding to a standardized mortality ratio (SMR) of 2.16 compared to age-matched controls^6^.

Despite these findings, there remains a paucity of data specifically addressing suicide risk in patients with CLL/SLL. Unlike patients with aggressive malignancies, those with CLL/SLL face a paradoxical combination of favorable long-term survival and enduring uncertainty about disease progression, therapeutic decision-making, and treatment-associated toxicities^7,8^. The chronic nature of CLL/SLL, coupled with the frequent adoption of a “watch-and-wait” management strategy, can lead to sustained psychological distress - often underrecognized and insufficiently addressed in clinical settings^8^.

Recent studies indicate that emotional distress among patients with CLL/SLL persists irrespective of disease stability, affecting both those undergoing active surveillance and those receiving treatment^9^. Despite this, routine mental health support remains limited, and validated predictive tools for suicide risk in this population are lacking. To address this unmet need, we analyzed a large, population-based dataset from the Surveillance, Epidemiology, and End Results (SEER) Program to assess suicide incidence and identify independent risk factors among CLL/SLL patients. We also developed a clinically practical nomogram to predict individual suicide risk. Our findings aim to enhance psychosocial screening in CLL/SLL survivors, support targeted mental health interventions, and ultimately improve survivorship care in this growing patient population.

## Materials and Methods

### Study Population and Data acquisition

Data for this study were obtained from the Surveillance, Epidemiology, and End Results (SEER) Program (https://seer.cancer.gov/), maintained by the National Cancer Institute (NCI) ^10^. The data were retrieved using SEER*Stat software version 8.4.3 (https://seer.cancer.gov/seerstat/, accessed on August 1, 2024) ^11^. Through the SEER 17 database [Incidence-SEER Research Data, 17 registries, Nov 2023 Sub (2000-2021)], patients diagnosed with CLL/SLL between 2000 and 2021 were extracted from the database using the “case listing session”. Only cases with known age (censored at age 89 years) and malignant behavior were included. The inclusion criteria were as follows: the International Classification of Diseases for Oncology (ICD-O-3) histologic code (9823/3). The exclusion criteria were as follows: (1) the diagnosis confirmation was “radiography without microscopic confirm” or “unknown”; (2) the patient’s survival time was 0 or unknown; (3) the sequence number was unknown; (4) age under 20 years at diagnosis.

Ultimately, 95,517 patients with CLL/SLL were included in the final cohort. As the SEER database is publicly available, deidentified and anonymized, ethical approval was not required. A flowchart illustrating the case selection process is presented in Figure 1.

**Figure 1.**
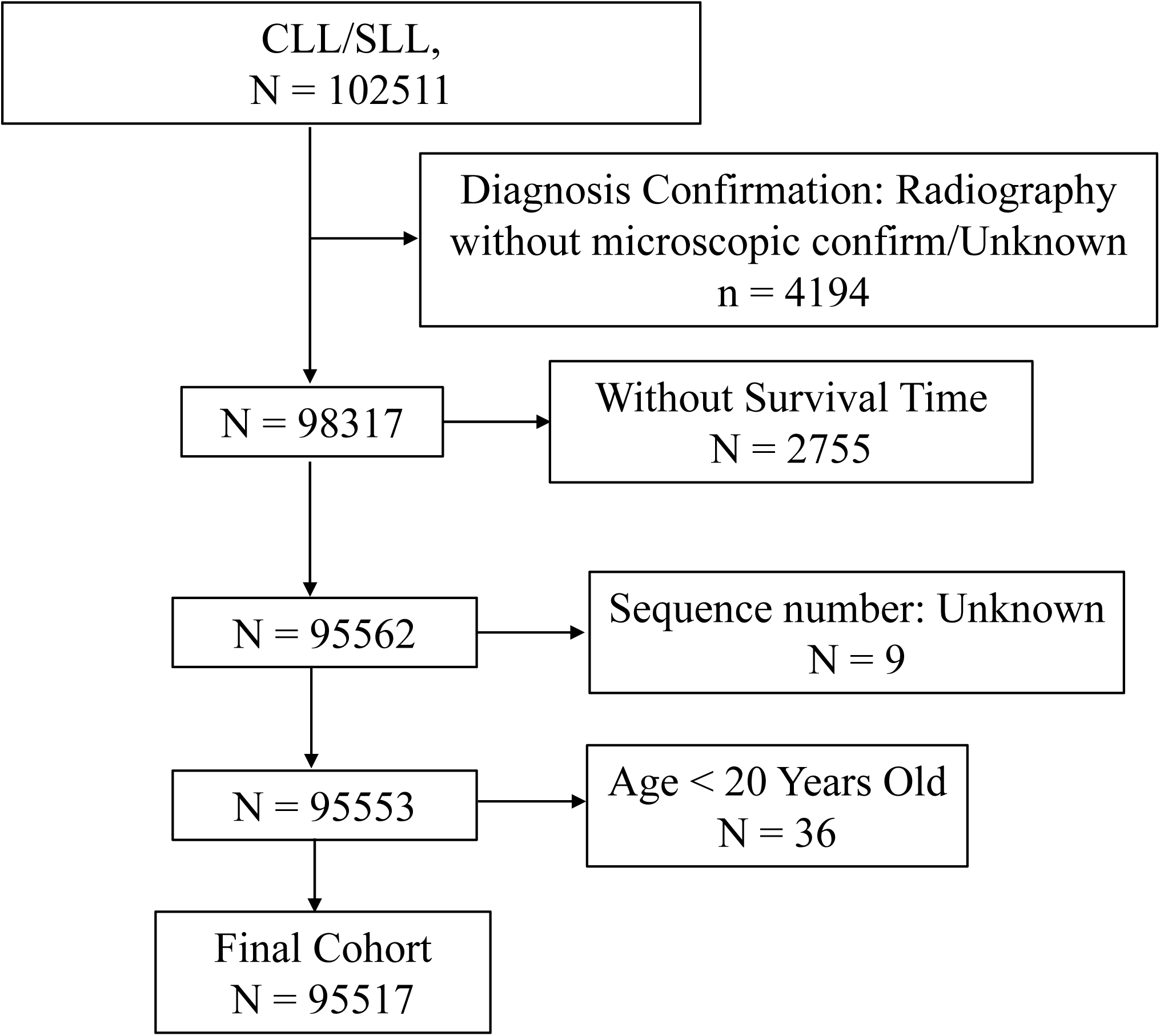
Flow chart of study cohort selection using the SEER database. A flow diagram of selection of patients with CLL/SLL in this study. CLL/SLL, Chronic lymphocytic leukemia/small lymphocytic lymphoma; SEER, Surveillance, Epidemiology, and End Results.

### Definition and Categorization of Study Variables

The analysis incorporated the following variables: age, sex, race, marital status, year of diagnosis, primary site, vital status, survival months, cause of death (COD) to site recode, cause-specific death classification, cause of death to site, sequence number, first malignant primary indicator, total number of in situ/malignant tumors for the patient, type of reporting source, diagnostic confirmation and chemotherapy recode. Age at diagnosis was categorized into four groups: 20-59, 60-69, 70-79 and ≥80 years old. Race was classified as White and Other (comprising “African American”, “Asian/Pacific Islander,” “American Indian/Alaska Native,” and “Unknown”). Marital status was grouped into married, single, or other (encompassing “divorced,” “separated,” “widowed,” “unmarried or domestic partner,” and “unknown”). Income was taken from the variable “Median Household Income in the past 12 months (in 2021 Inflation Adjusted Dollars)” and categorized as <$50,000, $50,000-$75,000, $75,000-$100,000 and $100,000+.

Residential status was divided based on urbanicity and population size into four categories: non-metropolitan areas, metropolitan areas with <250,000 people, 250,000 - 1 million people, and >1 million people. COD was defined using the “COD to site recode” variable. Diagnosis year was grouped into “2000-2004,” “2005-2009,” “2010-2015,” and “2016-2021.”

### Statistical analysis

All statistical analyses were conducted using the R program language (version 4.2.1; R Foundation for Statistical Computing, Vienna, Austria). Patients were randomly assigned to training and validation cohorts in a 7:3 ratio. Continuous variables were analyzed using Student’s *t*-test, while categorical variables were compared using *chi*-square test or Fisher’s exact test, as appropriate. Suicide risk factors for CLL/SLL were initially screened through least absolute shrinkage and selection operator (LASSO) regression, followed by univariate and multivariate Cox proportional regression analyses. To account for competing events, Fine-Gray competing risk regression was also conducted to identify independent predictors. Hazard ratios (HR) and 95% confidence intervals (95% CIs) were reported. The Cox proportional hazards regression model was assessed for proportionality assumptions, with no violations detected. Variables with *P* < 0.05 in all models were considered significant. Nomograms predicting 3-, 5- and 10-year suicide risk probabilities were constructed based on selected independent prognostic factors.

Model performance was evaluated using time-dependent receiver operating characteristic (ROC) curves, with the area under the curve (AUC) values indicating discriminatory ability. Calibration plots were used to assess agreement between predicted and observed probabilities. Decision curve analysis (DCA) was conducted to evaluate the net clinical benefit. CLL/SLL patients were stratified into high- and low-risk subgroups based on the median nomogram score. Competing risk analysis was then performed to estimate suicide risk across groups, and differences were assessed using Gray’s test. A two-sided *P* value < 0.05 indicated statistical significance.

## Results

### Baseline Characteristics of CLL/SLL Patients

As depicted in Figure 1, a total of 95,517 patients diagnosed with CLL/SLL were identified from the SEER 17 registry, Nov 2023 Sub (2000-2021) from January 2000 to December 2021. Among the study cohort, 60.2% were male, approximately 1.5 times the number of female patients (*n*=37,982, 39.8%; Table 1). The mean age at diagnosis was 69.2 ± 11.6 years. Age distribution was as follows: 20.7% were aged 20-59 years, 28.2% were 60-69 years, 29.5% were 70-79 years, and 21.6% were aged 80 years or older (≥80). Most CLL/SLL patients were white (88.8%), while the remaining 11.2% belonged to other racial group (including African American, Asian/Pacific Islander, American Indian/Alaska Native and “unknown”). At diagnosis, 54.4% of patients were married, 35.4% had other marital statuses (such as divorced, separated, widowed, unmarried or domestic partner), and 10.2% were single, having never been married. Among all identified cases, 79.9% were primary CLL/SLL and 20.1% were secondary CLL/SLL that were secondary to other primary malignancies. About 15.9% were treated with chemotherapy. At the time of the last follow-up, 522,94 (54.7%) patients were alive. A total of 17,840 (18.7%) patients had died due to CLL/SLL, while 138 (0.1%) patients died of suicide, and an additional 25,245 (26.4%) deaths were attributed to other causes, including cerebrovascular diseases, lung diseases, septicemia, and so on. There were no statistically significant differences in these baseline characteristics between the training and validation cohorts (*P* > 0.05). A detailed comparison of epidemiologic features was summarized in Table 1.

### LASSO Regression and Selection of Independent Prognostic Factors

A total of 10 clinical variables were included in the training cohort. Based on the results of LASSO regression analysis, except for sequence and treatment delay, the other 8 variables including age, sex, race, diagnosis year, marital status at diagnosis, median household income, residence type and chemotherapy were identified as relevant suicide risk factors by using the minimum standard value as the criterion (Figure 2). The Cox regression model was further used to screen the prognostic factors. All the 8 variables passed the preliminary proportional hazards assumption test. Univariate Cox regression analysis revealed that age, sex, race, marital status at diagnosis, and income were significantly associated with suicide risk (Table 2). In the multivariate Cox analysis, age, sex, race, marital status at diagnosis, and income were also independently and significantly associated with suicide risk (Table 2). Aged over 80 years (HR 2.44, 95% CI: 1.35-4.42, *P* = 0.003), marital status of single (HR 2.35, 95% CI: 1.25-4.40, *P* = 0.008) and marital status of other (including divorced, widowed, separated, unmarried or domestic partner and unknown at diagnosis; HR 2.44, 95% CI: 1.59-3.73, *P* < 0.001) were significantly associated with increased suicide risk, while gender of female (HR 0.30, 95% CI: 0.06-0.24, *P* < 0.001), race of other (races of not white; HR 0.33, 95% CI: 0.12-0.91, *P* = 0.032) and median household income over $100,000 (in 2021 Inflation Adjusted Dollars; HR 0.30, 95% CI: 0.12-0.72, *P* = 0.007) were remarkably associated with decreased suicide risk. The detailed results were demonstrated in Table 2. To account for competing risks, Fine-Gray modeling was employed. This analysis confirmed that sex, race, marital status at diagnosis, and income remained independent predictors of suicide risk, while age was no longer statistically significant. These four variables were subsequently selected to construct a nomogram for predicting the 3-, 5- and 10-year suicide probability of CLL/SLL patients (Figure 3).

**Figure 2.**
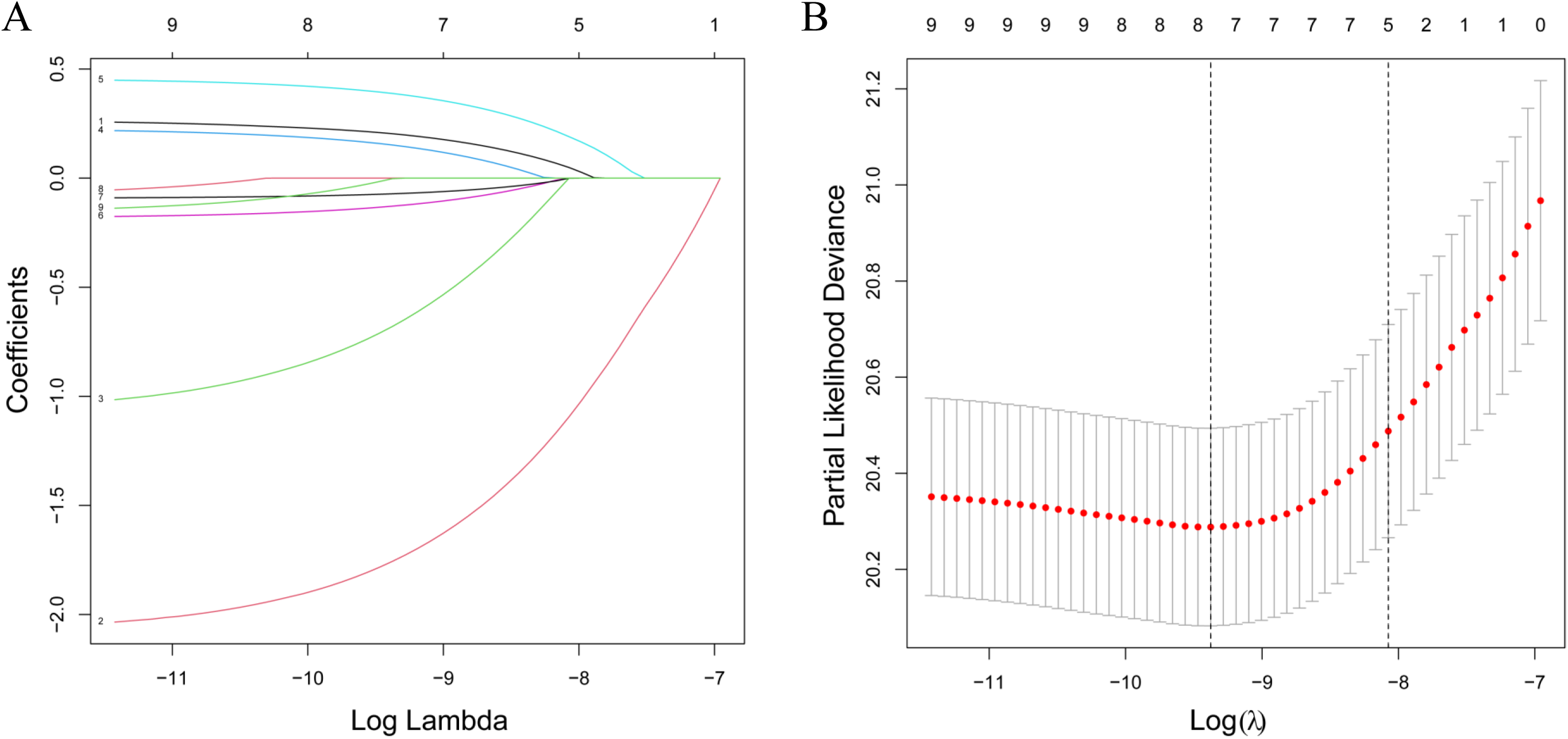
Refining predictive factors through Least Absolute Shrinkage and Selection Operator (LASSO) regression model. (A) LASSO coefficients of 10 clinical features; (B) Selection of the optimum parameter (λ) for LASSO model.

**Figure 3.**
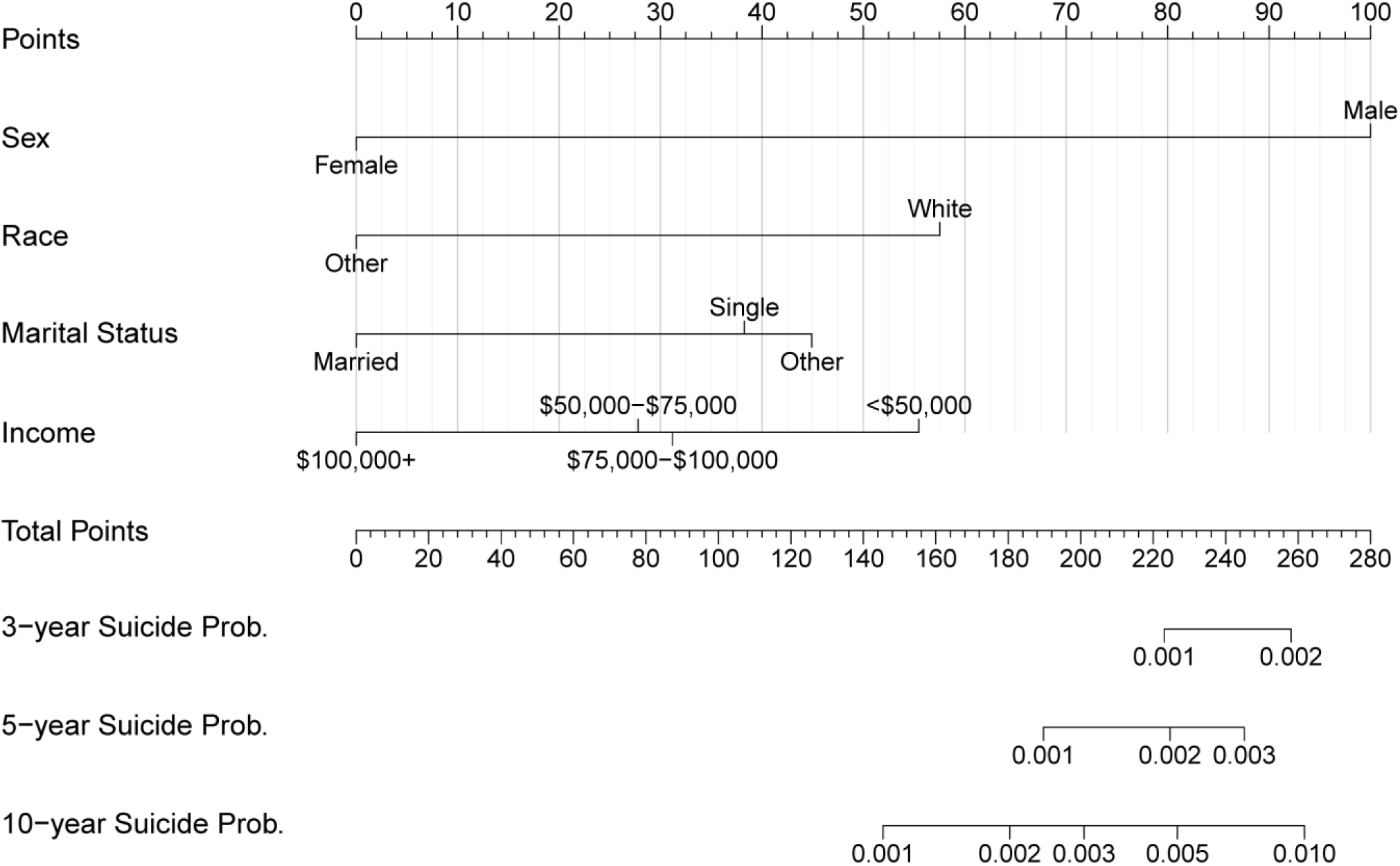
Construction of the prognostic nomogram to predict the 3-, 5-, and 10-year suicide risk in CLL/SLL patients. The total points were calculated by integrating scores related to sex, race, marital status and income, and were then projected to the bottom scale to predict the suicide probability at 3, 5 and 10 years, respectively. CLL/SLL, Chronic lymphocytic leukemia/small lymphocytic lymphoma.

### Performance Evaluation and Validation of the Suicide Risk Nomogram

The predictive performance of the suicide risk nomogram was assessed using time-dependent ROC curves and calibration plots in both the training and validation cohorts. Time-dependent ROC analyses demonstrated that the nomogram exhibited remarkable accuracy for predicting suicide risk at 3, 5, and 10 years, with AUC values of 0.786, 0.735, and 0.734, respectively, in the training cohort (Figure 4A), and 0.711, 0.739, and 0.727, respectively, in the validation cohort (Figure 4B). The calibration curves showed good agreement between prediction and actual observation at the 3-, 5-, and 10-year intervals in both the training cohort (Figure 5A-C) and the validation cohort (Figure 5D-F). DCA was further employed to evaluate the clinical net benefit of the predictive nomogram model. The results showed that the nomogram model has a favorable net benefit in predicting the 3-, 5- and 10-year suicide risk in both the training (Figure 6 A-C) and validation cohorts (Figure 6D-F).

**Figure 4.**
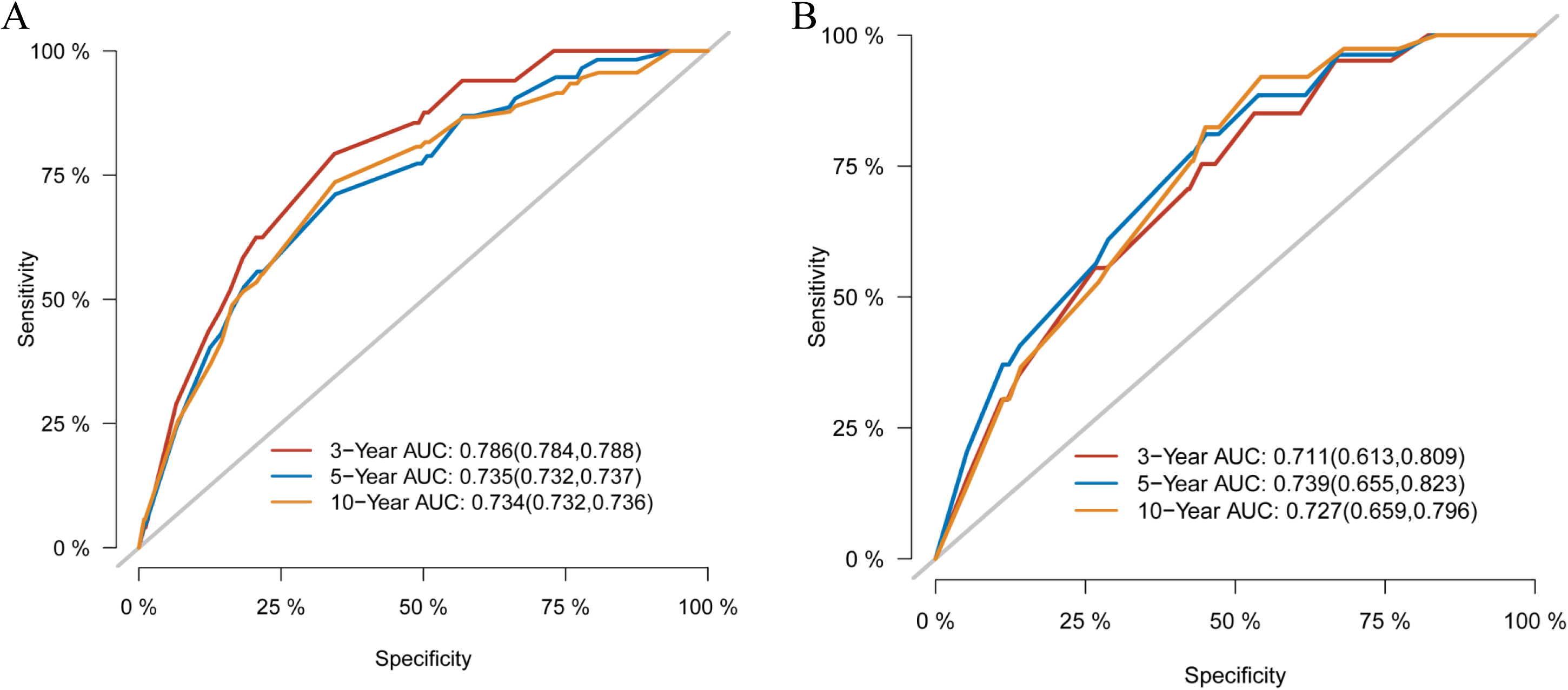
Evaluation of the nomogram by time-dependent receiver operating characteristic (ROC) curves in the training cohort (A) and validation cohort (B).

**Figure 5.**
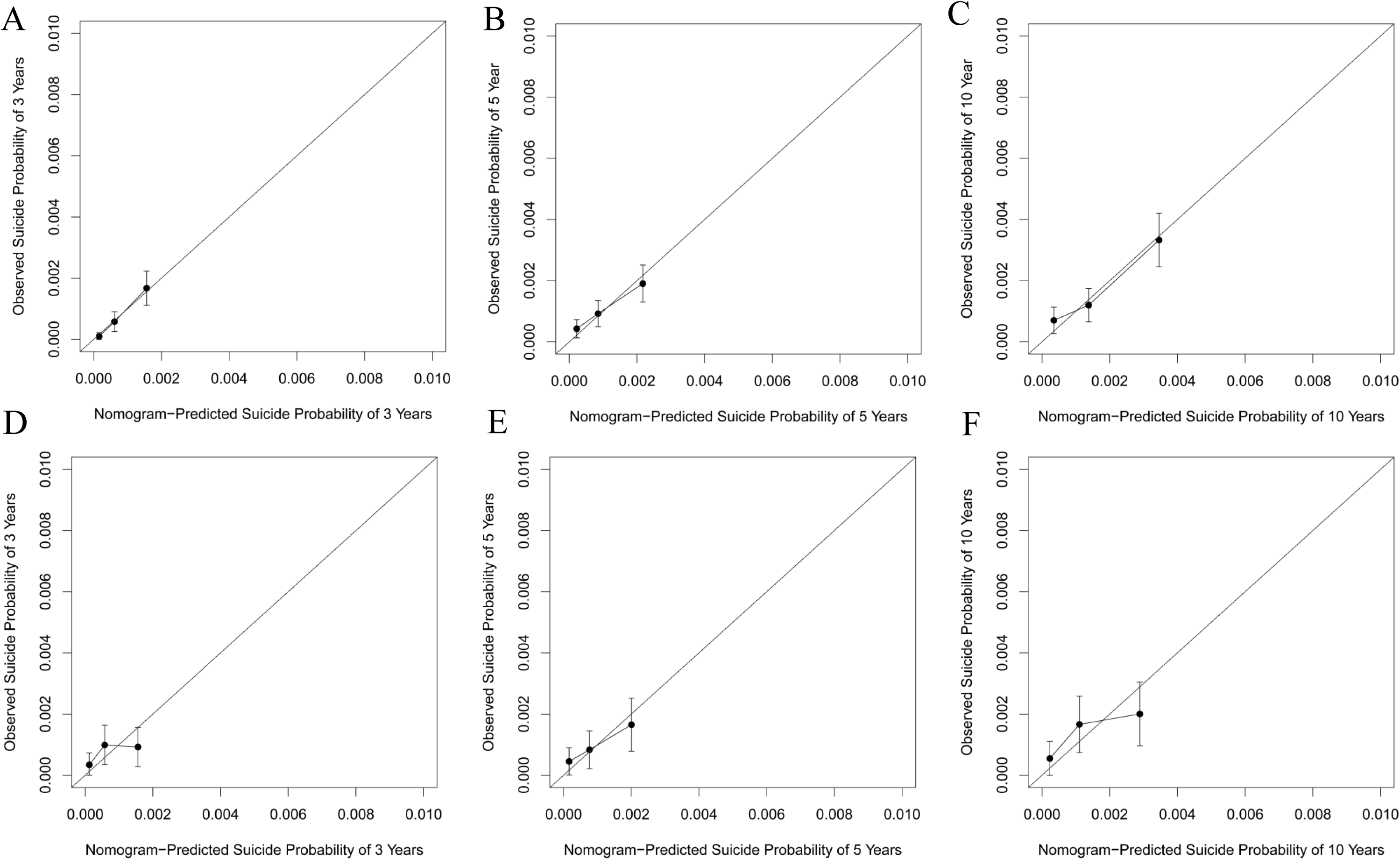
Calibration plots for nomogram predicting suicide risk in CLL/SLL patients. (A-C) The calibration curves of the training cohort for the observed overall suicide probability and predicted suicide probability at 3 years, 5 years and 10 years, respectively. (D-F) The calibration curves of the validation cohort at 3 years, 5 years and 10 years, respectively. CLL/SLL, Chronic lymphocytic leukemia/small lymphocytic lymphoma.

**Figure 6.**
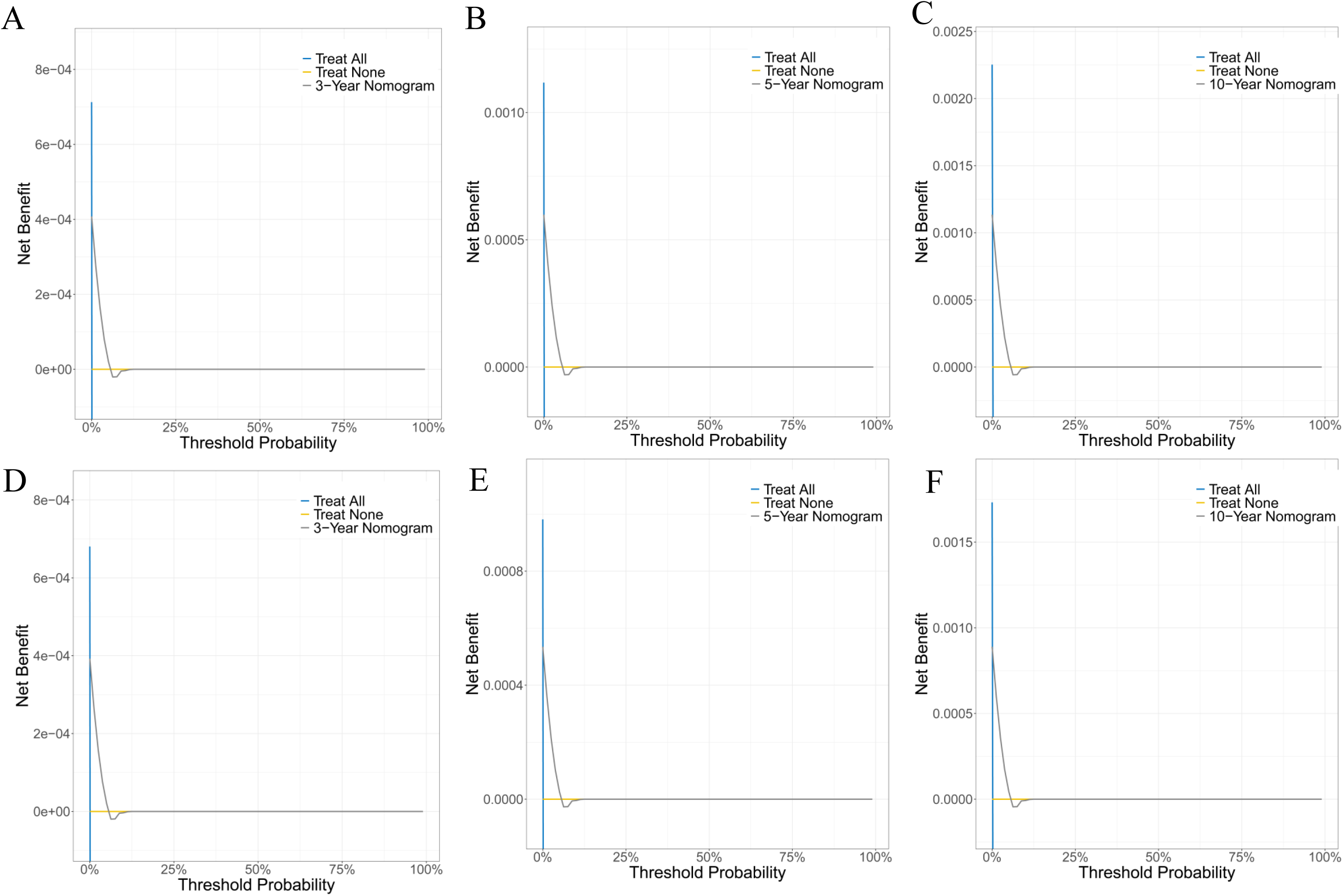
Evaluation of the nomogram of by decision curve analysis. (A-C) The decision curve analysis of the nomogram at 3 years (A), 5 years (B) and 10 years (C) in the training cohort. (D-F) The decision curve analysis of the nomogram at 3 years (D), 5 years (E) and 10 years (F) in the validation cohort.

### Competing Risk Analysis Between the Stratified Risk Groups

Risk scores for each variable were derived from the nomogram, and total cumulative scores were subsequently calculated for all patients. Based on the median risk score, the entire cohort was stratified into low- and high-risk subgroups. Competing risk analysis demonstrated significant differences in cumulative suicide incidence between the low- and high-risk groups in both the training cohort (Gray’s test, *P* < 0.001; Figure 7A) and the validation cohort (Gray’s test, *P* < 0.001; Figure 7B), highlighting the nomogram’s robust capability for effective risk stratification.

**Figure 7.**
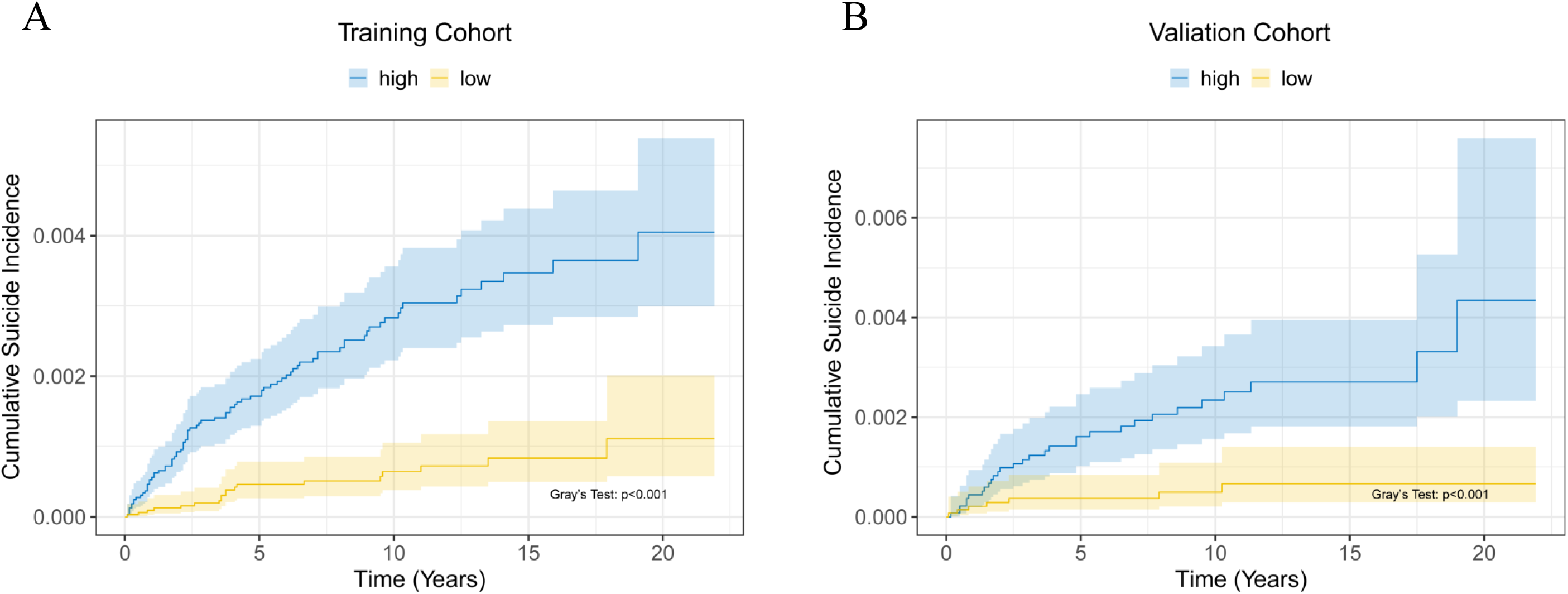
The cumulative suicide incidence of CLL/SLL patients stratified by risk stratification using the constructed nomogram in the training cohort (A) and validation cohort (B). CLL/SLL, Chronic lymphocytic leukemia/small lymphocytic lymphoma.

## Discussion

This large population-based analysis of 95,517 patients with CLL/SLL from the SEER 17 registry (2000-2021) provides important insights into suicide risk in this patient population. To our knowledge, this is the most comprehensive study to date assessing independent risk factors for suicide among CLL/SLL patients and establishing a validated predictive model to quantify suicide risk over time. The baseline characteristics of the cohort reaffirm that CLL/SLL predominantly affects older adults, with a mean age at diagnosis of 69.2 years and a substantial proportion (21.6%) aged ≥80 years. The male-to-female ratio of approximately 1.5:1, combined with a high predominance of white patients (88.8%), is consistent with known epidemiologic patterns in these indolent B-cell malignancies^1,12,13^.

Consistent with patterns reported in other hematologic malignancies^6,14,15^, our study found that suicide, although relatively rare (0.1%), represents a clinically significant cause of mortality in patients with CLL/SLL, particularly when compared to the general population. The multivariate Cox and Fine-Gray competing risk analyses revealed that several sociodemographic variables, namely age, sex, race, marital status, and income, are independently associated with suicide risk. Advanced age (≥80 years) was a strong predictor of suicide in the multivariate Cox analysis (HR 2.44), echoing prior findings that older patients with cancer experience heightened psychological vulnerability, often compounded by comorbidities, functional decline, and social isolation^16,17^. However, age lost statistical significance in the competing risk model, suggesting that its effect may be mediated or confounded by other factors when considering non-suicide mortality as a competing risk. In contrast, male sex emerged as a consistently significant risk factor in all analyses, with men showing more than threefold higher risk of suicide than women (HR 0.30 for female sex), aligning with well-established trends in oncology and general populations^4,18,19^.

Notably, marital status at diagnosis was a strong and independent predictor of suicide^16^. In this study, patients who were single (never married) or in non-married categories (divorced, widowed, separated, or domestic partner) demonstrated over twice the risk of suicide compared to their married counterparts. These findings support the protective effect of social support, particularly spousal or family relationships, as documented in previous oncologic studies^15,16^. Similarly, socioeconomic status, represented by median household income, was inversely associated with suicide risk. Patients with median household income above $100,000 (USD, inflation-adjusted) had a significantly lower risk of suicide (HR 0.30), reinforcing the detrimental impact of financial hardship and health-related economic stress^4^.

Our analysis also demonstrated that patients identifying as non-White had significantly lower suicide risk compared to White patients (HR 0.33), a finding consistent with national epidemiological data^15,18,20^. While these differences may reflect cultural, social, or reporting variations in mental health and suicide, they underscore the importance of considering race-specific vulnerabilities and resilience factors in future psycho-oncological research^15^.

Importantly, using the four most stable predictors: sex, race, marital status, and income, we developed and validated a nomogram capable of estimating individualized suicide risk at 3, 5, and 10 years^21^. The nomogram exhibited strong discriminative ability, with AUC values exceeding 0.71 in both the training and validation cohorts. Calibration curves confirmed that predicted probabilities closely matched observed outcomes across time intervals, and decision curve analyses demonstrated a favorable net clinical benefit, reinforcing its potential utility in real-world settings^22^.

The stratification of patients into high- and low-risk groups based on the nomogram scores further confirmed its prognostic robustness, as evidenced by significantly different cumulative suicide incidences between groups in both training and validation cohorts. This stratification tool may serve as a valuable resource for clinicians aiming to identify vulnerable patients early in the disease course and integrate psychosocial support more effectively into CLL/SLL management^20,23^.

However, our study’s findings must be contextualized within the broader literature on suicide risk in cancer. Multiple reports have documented an elevated risk of suicide among patients with hematologic malignancies, often attributing this risk to a combination of chronic psychological distress, social isolation, and economic hardship^6,14,20^. In CLL/SLL, the “watch-and-wait” strategy combined with a protracted disease course likely exacerbates these stressors, even in the context of relatively favorable survival outcomes^8,23^. The independent predictive strength of marital status and income in our analysis underscores the importance of social support systems and financial security as buffers against psychological distress^20^. Similarly, our finding of a protective effect in females and in patients of non-white race (when compared to white patients) is consistent with prior research suggesting that differences in coping styles and social support networks may modulate suicide risk^6,16^.

Despite the strengths of our study, including its large sample size and long follow-up period, the relatively low number of suicide events (0.1%) poses inherent challenges for risk modeling. Moreover, certain limitations should be acknowledged. First, the SEER database lacks detailed clinical data on disease stage, performance status, comorbid psychiatric diagnoses, and substance use, which may further refine suicide risk assessment. Additionally, suicide may be underreported in registry data, potentially leading to conservative estimates of incidence. Lastly, while our nomogram demonstrated strong internal validity, external validation in other populations with diverse demographic and healthcare backgrounds is warranted before broad clinical implementation.

## Conclusions

In conclusion, this study provides robust evidence that psychosocial factors, specifically marital status, income, gender, and race, are key determinants of suicide risk in CLL/SLL patients. Our predictive nomogram, rooted in these independent risk factors, offers a clinically applicable tool to facilitate suicide risk prediction and aid in early identification of vulnerable patients. By integrating such risk stratification methods into routine clinical practice, oncologists and hematologists may be better equipped to provide targeted psychosocial interventions and ultimately improve survivorship care outcomes in this growing patient population.

## Data Availability

The data analyzed in this study are from the SEER database (https://seer.cancer.gov/) that are available to the public.

## Acknowledgments

The interpretation of the data is the sole responsibility of the author(s). The author(s) acknowledge the efforts of the National Cancer Institute and the Surveillance, Epidemiology, and End Results (SEER) Program tumor registries in the creation of the SEER database.

## Abbreviations

CLL/SLL: Chronic lymphocytic leukemia/small lymphocytic lymphoma
Chemo: chemotherapy
CI: confidence interval
COD: cause of death
DCA: decision curve analysis
HR: hazard ration
OS: overall survival
ROC: receiver operating characteristic
SEER: Surveillance, Epidemiology, and End Results.

## References

1. Jain N, Wierda WG, O’Brien S. Chronic lymphocytic leukaemia. The Lancet. 2024;404(10453):694–706. doi:10.1016/S0140-6736(24)00595-6

2. Shadman M. Diagnosis and Treatment of Chronic Lymphocytic Leukemia: A Review. JAMA. 2023;329(11):918–932. doi:10.1001/jama.2023.1946

3. Tam C, Thompson PA. BTK inhibitors in CLL: second-generation drugs and beyond. Blood Advances. 2024;8(9):2300–2309. doi:10.1182/bloodadvances.2023012221

4. Zaorsky NG, Zhang Y, Tuanquin L, Bluethmann SM, Park HS, Chinchilli VM. Suicide among cancer patients. Nature Communications. 2019/01/14 2019;10(1):207. doi:10.1038/s41467-018-08170-1

5. Henson KE, Brock R, Charnock J, Wickramasinghe B, Will O, Pitman A. Risk of Suicide After Cancer Diagnosis in England. JAMA Psychiatry. 2019;76(1):51–60. doi:10.1001/jamapsychiatry.2018.3181

6. Yu H, Cai K, Huang Y, Lyu J. Risk factors associated with suicide among leukemia patients: A Surveillance, Epidemiology, and End Results analysis. Cancer Medicine. 2020;9(23):9006–9017. 10.1002/cam4.3502

7. Landau DA, Sun C, Rosebrock D, et al. The evolutionary landscape of chronic lymphocytic leukemia treated with ibrutinib targeted therapy. Nature Communications. 2017/12/19 2017;8(1):2185. doi:10.1038/s41467-017-02329-y

8. Hallek M. Chronic Lymphocytic Leukemia: 2025 Update on the Epidemiology, Pathogenesis, Diagnosis, and Therapy. American Journal of Hematology. 2025;100(3):450–480. 10.1002/ajh.27546

9. Levin TT, Li Y, Riskind J, Rai K. Depression, anxiety and quality of life in a chronic lymphocytic leukemia cohort. General Hospital Psychiatry. 2007/05/01/ 2007;29(3):251–256. 10.1016/j.genhosppsych.2007.01.014

10. Surveillance, Epidemiology, and End Results (SEER) Program (www.seer.cancer.gov) SEER*Stat Database: Incidence - SEER Research Plus Data, 17 Registries, Nov 2022 Sub (2000-2020) - Linked To County Attributes - Time Dependent (1990-2021) Income/Rurality, 1969-2021 Counties, National Cancer Institute, DCCPS, Surveillance Research Program, released April 2023, based on the November 2022 submission.

11. Surveillance Research Program, National Cancer Institute SEER*Stat software (seer.cancer.gov/seerstat) version 8.4.3.

12. Alaggio R, Amador C, Anagnostopoulos I, et al. The 5th edition of the World Health Organization Classification of Haematolymphoid Tumours: Lymphoid Neoplasms. Leukemia. 2022/07/01 2022;36(7):1720–1748. doi:10.1038/s41375-022-01620-2

13. Lumish M, Falchi L, Imber BS, Scordo M, von Keudell G, Joffe E. How we treat mature B-cell neoplasms (indolent B-cell lymphomas). Journal of Hematology & Oncology. 2021/01/06 2021;14(1):5. doi:10.1186/s13045-020-01018-6

14. Shen J, Lin S, Tao H, Sechi LA, Fozza C, Wen X. Risk assessment and predictive modeling of suicide in multiple myeloma patients. Journal of Cancer Survivorship. 2024/12/17 2024;doi:10.1007/s11764-024-01732-x

15. Zhou J, Tian M, Zhang X, et al. Suicide among lymphoma patients. Journal of Affective Disorders. 2024/09/01/ 2024;360:97–107. 10.1016/j.jad.2024.05.158

16. Liu Q, Wang X, Kong X, et al. Subsequent risk of suicide among 9,300,812 cancer survivors in US: A population-based cohort study covering 40 years of data. eClinicalMedicine. 2022;44doi:10.1016/j.eclinm.2022.101295

17. Hu X, Ma J, Jemal A, et al. Suicide Risk Among Individuals Diagnosed With Cancer in the US, 2000-2016. JAMA Network Open. 2023;6(1):e2251863–e2251863. doi:10.1001/jamanetworkopen.2022.51863

18. Han X, Hu X, Zhao J, Ma J, Jemal A, Yabroff KR. Trends of Cancer-Related Suicide in the United States: 1999-2018. JNCI: Journal of the National Cancer Institute. 2021;113(9):1258–1262. doi:10.1093/jnci/djaa183

19. Guo Z, Gu C, Li S, et al. Incidence and risk factors of suicide among patients diagnosed with bladder cancer: A systematic review and meta-analysis. Urologic Oncology: Seminars and Original Investigations. 2021/03/01/ 2021;39(3):171–179. 10.1016/j.urolonc.2020.11.022

20. Kim J, Gang M. Identifying the Risk Factors of Suicidal Ideation in Patients With Hematologic Malignancies Using a Multidimensional Approach. Western Journal of Nursing Research. 2024;46(2):81–89. doi:10.1177/01939459231216870

21. Balachandran VP, Gonen M, Smith JJ, DeMatteo RP. Nomograms in oncology: more than meets the eye. The Lancet Oncology. 2015;16(4):e173–e180. doi:10.1016/S1470-2045(14)71116-7

22. Zheng S, Tong Y, Chen J, Yang L, Tan Y. Construction and evaluation of leukemia suicide risk predictive model based on SEER database. Original Research. Frontiers in Psychiatry. 2025-February-21 2025;Volume 16 - 2025doi:10.3389/fpsyt.2025.1506550

23. Fedele PL, Opat S. Chronic Lymphocytic Leukemia: Time to Care for the Survivors. Journal of Clinical Oncology. 2024;42(17):2005–2011. doi:10.1200/jco.23.02738

