## Supplementary material for "Risk Assessment and Predictive Modeling of Suicide in Chronic Lymphocytic Leukemia/Small Lymphocytic Lymphoma (CLL/SLL) Patients": Table 1

**Table 1** Comparison of baseline characteristics of CLL/SLL patients between the training set and validation set from SEER database.

| **Characteristics** | **Total (N=95517)** | **Training cohort (N=66885)** | **Validation cohort (N=28632)** | ***P*-value** |
| --- | --- | --- | --- | --- |
| **Age*, n (%)*** |  |  |  |  |
| 20-59 | 19773 (20.7%) | 13811 (20.6%) | 5962 (20.8%) | 0.219 |
| 60-69 | 26981 (28.2%) | 18798 (28.1%) | 8183 (28.6%) |  |
| 70-79 | 28160 (29.5%) | 19747 (29.5%) | 8413 (29.4%) |  |
| ≥80 | 20603 (21.6%) | 14529 (21.7%) | 6074 (21.2%) |  |
| **Sex*, n (%)*** |  |  |  |  |
| Male | 57535 (60.2%) | 40381 (60.4%) | 17154 (59.9%) | 0.184 |
| Female | 37982 (39.8%) | 26504 (39.6%) | 11478 (40.1%) |  |
| **Race*, n (%)*** |  |  |  |  |
| White | 84789 (88.8%) | 59353 (88.7%) | 25436 (88.8%) | 0.666 |
| Other^1^ | 10728 (11.2%) | 7532 (11.3%) | 3196 (11.2%) |  |
| **Diagnosis Year*, n (%)*** |  |  |  |  |
| 2000-2004 | 15453 (16.2%) | 10841 (16.2%) | 4612 (16.1%) | 0.479 |
| 2005-2009 | 17962 (18.8%) | 12494 (18.7%) | 5468 (19.1%) |  |
| 2010-2015 | 30443 (31.9%) | 21324 (31.9%) | 9119 (31.8%) |  |
| 2016-2021 | 31659 (33.1%) | 22226 (33.2%) | 9433 (32.9%) |  |
| **Sequence*, n (%)*** |  |  |  |  |
| Primary CLL/SLL^2^ | 76318 (79.9%) | 53550 (80.1%) | 22768 (79.5%) | 0.056 |
| Secondary CLL/SLL^3^ | 19199 (20.1%) | 13335 (19.9%) | 5864 (20.5%) |  |
| **Marital Status*, n (%)*** |  |  |  |  |
| Married | 51993 (54.4%) | 36449 (54.5%) | 15544 (54.3%) | 0.443 |
| Single^4^ | 9720 (10.2%) | 6752 (10.1%) | 2968 (10.4%) |  |
| Other^5^ | 33804 (35.4%) | 23684 (35.4%) | 10120 (35.3%) |  |
| **Income**^6^***, n (%)*** |  |  |  |  |
| <$50,000 | 5967 (6.2%) | 4152 (6.2%) | 1815 (6.3%) | 0.491 |
| $50,000-$75,000 | 36144 (37.8%) | 25404 (38.0%) | 10740 (37.5%) |  |
| $75,000-$100,000 | 35940 (37.6%) | 25095 (37.5%) | 10845 (37.9%) |  |
| $100,000+ | 17466 (18.3%) | 12234 (18.3%) | 5232 (18.3%) |  |
| **Residence*, n (%)*** |  |  |  |  |
| Nonmetropolitan | 13020 (13.6%) | 9185 (13.7%) | 3835 (13.4%) | 0.175 |
| Metropolitan < 250,000^7^ | 8335 (8.7%) | 5840 (8.7%) | 2495 (8.7%) |  |
| Metropolitan 250,000 - 1 million^8^ | 19828 (20.8%) | 13960 (20.9%) | 5868 (20.5%) |  |
| Metropolitan > 1 million^9^ | 54334 (56.9%) | 37900 (56.7%) | 16434 (57.4%) |  |
| **Treatment Delay (days)*, n (%)*** |  |  |  |  |
| ≤30 | 8257 (8.6%) | 5742 (8.6%) | 2515 (8.8%) | 0.659 |
| 31-90 | 4427 (4.6%) | 3082 (4.6%) | 1345 (4.7%) |  |
| 91-120 | 738 (0.8%) | 522 (0.8%) | 216 (0.8%) |  |
| >120 | 82095 (85.9%) | 57539 (86.0%) | 24556 (85.8%) |  |
| **Chemo*, n (%)*** |  |  |  |  |
| No/Unknown | 80336 (84.1%) | 56349 (84.2%) | 23987 (83.8%) | 0.070 |
| Yes | 15181 (15.9%) | 10536 (15.8%) | 4645 (16.2%) |  |
| **COD*, n (%)*** |  |  |  |  |
| Alive | 52294 (54.7%) | 36539 (54.6%) | 15755 (55.0%) | 0.562 |
| CLL/SLL^10^ | 17840 (18.7%) | 12544 (18.8%) | 5296 (18.5%) |  |
| Suicide^11^ | 138 (0.1%) | 101 (0.2%) | 37 (0.1%) |  |
| Other cause^12^ | 25245 (26.4%) | 17701 (26.5%) | 7544 (26.3%) |  |

^1^ Races including African American, Asian/Pacific Islander, American Indian/Alaska Native and “unknown”.

^2^ Sequence number of “primary only” and “1st of 2 or more primaries”, indicating CLL/SLL was the primary malignancy.

^3^ Sequence number of “2nd of 2 or more primaries”, “3rd of 3 or more primaries” and “4th of 4 or more primaries”, indicating CLL/SLL was secondary to other primary malignancies.

^4^ Marital status at diagnosis was single (never married).

^5^ Marital statuses of divorced, widowed, separated, unmarried or domestic partner and unknown at diagnosis.

^6^ Median Household Income in the past 12 months (in 2021 Inflation Adjusted Dollars).

^7^ Living in a metropolitan area with a population of less than 250,000.

^8^ Living in a metropolitan area with a population of about 250,000 to 1 million.

^9^ Living in a metropolitan area with a population of 1 million or more.

^10^ Death attributable to CLL/SLL.

^11^ Death due to suicide.

^12^ Dead of other cause such as diseases of heart, lung disease, septicemia, and so on.

Abbreviations: CLL/SLL, Chronic lymphocytic leukemia/small lymphocytic lymphoma; Chemo, chemotherapy; COD, cause of death.
