## Supplementary material for "Risk Assessment and Predictive Modeling of Suicide in Chronic Lymphocytic Leukemia/Small Lymphocytic Lymphoma (CLL/SLL) Patients": Table 2

**Table 2** Univariable and multivariable cox regression analysis and Fine-gray competing risks regression

model of Suicide Risk in the Training Cohort of CLL/SLL patients.

| **Parameters** | **Univariate Cox** | | **Multivariate Cox** | | **Fine-gray model** | |
| --- | --- | --- | --- | --- | --- | --- |
|  | **HR (95%CI)** | ***P*-value** | **HR (95%CI)** | ***P*-value** | **HR (95%CI)** | ***P*-value** |
| **Age** |  |  |  |  |  |  |
| 20-59 | Reference |  | Reference |  | Reference |  |
| 60-69 | 0.95(0.55-1.66) | 0.865 | 0.99(0.57-1.73) | 0.969 | 0.84(0.53-1.33) | 0.530 |
| 70-79 | 1.10(0.63-1.93) | 0.732 | 1.23(0.69-2.16) | 0.483 | 0.81(0.51-1.30) | 0.470 |
| ≥80 | 2.15(1.21-3.84) | 0.009 | 2.44(1.35-4.42) | 0.003 | 1.05(0.65-1.69) | 0.870 |
| **Sex** |  |  |  |  |  |  |
| Male | Reference |  | Reference |  | Reference |  |
| Female | 0.16 (0.08-0.31) | <0.001 | 0.12(0.06-0.24) | <0.001 | 0.14(0.08-0.25) | <0.001 |
| **Race** |  |  |  |  |  |  |
| White | Reference |  | Reference |  | Reference |  |
| Other^1^ | 0.36(0.13-0.98) | 0.045 | 0.33(0.12-0.91) | 0.032 | 0.31(0.13-0.72) | 0.022 |
| **Diagnosis Year** |  |  |  |  |  |  |
| 2000-2004 | Reference |  |  |  |  |  |
| 2005-2009 | 1.08(0.57-2.06) | 0.808 |  |  |  |  |
| 2010-2015 | 1.75(0.96-3.16) | 0.066 |  |  |  |  |
| 2016-2021 | 1.72(0.85-3.51) | 0.133 |  |  |  |  |
| **Sequence** |  |  |  |  |  |  |
| Primary CLL/SLL^2^ | Reference |  |  |  |  |  |
| Secondary CLL/SLL^3^ | 1.11(0.66-1.85) | 0.693 |  |  |  |  |
| **Marital Status** |  |  |  |  |  |  |
| Married | Reference |  | Reference |  | Reference |  |
| Single^4^ | 1.87(1.00-3.48) | 0.049 | 2.35(1.25-4.40) | 0.008 | 2.02(1.19-3.44) | 0.028 |
| Other^5^ | 1.86(1.22-2.82) | 0.004 | 2.44(1.59-3.73) | <0.001 | 2.30(1.60-3.30) | <0.001 |
| **Income**^6^ |  |  |  |  |  |  |
| <$50,000 | Reference |  | Reference |  | Reference |  |
| $50,000-$75,000 | 0.58(0.29-1.16) | 0.123 | 0.56(0.28-1.13) | 0.105 | 0.61(0.34-1.10) | 0.170 |
| $75,000-$100,000 | 0.62(0.31-1.24) | 0.174 | 0.58(0.29-1.16) | 0.124 | 0.66(0.37-1.17) | 0.230 |
| $100,000+ | 0.32(0.13-0.76) | 0.010 | 0.30(0.12-0.72) | 0.007 | 0.35(0.17-0.73) | 0.019 |
| **Residence** |  |  |  |  |  |  |
| Nonmetropolitan | Reference |  |  |  |  |  |
| Metropolitan < 250,000^7^ | 0.88(0.42-1.85) | 0.741 |  |  |  |  |
| Metropolitan 250,000 - 1 million^8^ | 0.75(0.40-1.38) | 0.354 |  |  |  |  |
| Metropolitan > 1 million^9^ | 0.60(0.35-1.01) | 0.055 |  |  |  |  |

^1^ Races including African American, Asian/Pacific Islander, American Indian/Alaska Native and “unknown”.

^2^ Sequence number of “primary only” and “1st of 2 or more primaries”, indicating CLL/SLL was the primary malignancy.

^3^ Sequence number of “2nd of 2 or more primaries”, “3rd of 3 or more primaries” and “4th of 4 or more primaries”, indicating CLL/SLL was secondary to other primary malignancies.

^4^ Marital status at diagnosis was single (never married).

^5^ Marital statuses of divorced, widowed, separated, unmarried or domestic partner and unknown at diagnosis.

^6^ Median Household Income in the past 12 months (in 2021 Inflation Adjusted Dollars).

^7^ Living in a metropolitan area with a population of less than 250,000.

^8^ Living in a metropolitan area with a population of about 250,000 to 1 million.

^9^ Living in a metropolitan area with a population of 1 million or more.

Abbreviations: CLL/SLL, Chronic lymphocytic leukemia/small lymphocytic lymphoma; Chemo, chemotherapy.
